# Racial differences in vaccine acceptance in a rural southern US state

**DOI:** 10.1101/2022.05.12.22274953

**Authors:** Benjamin C. Amick, Jaimi L. Allen, Clare C. Brown, Anthony Goudie, Mick Tilford, Mark Williams

## Abstract

**Introduction:** To assess vaccine acceptance among adults living in a largely rural Southern state.

**Methods:** Data were collected between October 3 and October 17, 2020 using random digit dialing. Participants included residents aged 18+, able to understand English or Spanish, and provide informed consent. The primary outcome was a multi-dimensional COVID-19 vaccine acceptance measure. Scores varied between -3 to +3.

**Results:** The sample (n=1,164) was weighted to be representative of the state’s population. Black participants had the lowest overall vaccine acceptance (0.5) compared to White participants (1.2). Hispanic participants had the highest scores (1.4). In adjusted models, Black participants had 0.81 points lower acceptance than White participants, and Hispanic participants had 0.35 points higher acceptance. Hispanic participants had the highest scores for all five vaccine acceptance dimensions, relatively equivalent to White participants. Black participants had consistently lower scores, especially perceived vaccine safety (mean -0.2, SD 0.1).

**Conclusions:** The lowest vaccine acceptance rates were among Black participants particularly on perceived vaccine safety. While Black participants had the lowest acceptance scores, Hispanic participants had the highest. This variability shows the value of a multi-dimensional vaccine acceptance measure to inform COVID-19 vaccination campaign strategies.

## Introduction

More than 34 million Americans have contracted COVID-19 and over 609,000 have died from the disease as of July 2021.^1^ Three vaccines have received emergency use authorization from the FDA and have been distributed throughout the United States. While over 49% of Americans have been fully vaccinated,^1^ less than 36% of the population in Arkansas have been fully vaccinated.^2^ Even with effective COVID-19 vaccines, there is little expectation Americans will accept the vaccine in sufficient numbers to achieve herd immunity.^3-4^ Mistrust and doubts plaguing vaccinations in general^5^ prevent the acceptance of a COVID-19 vaccine.^6^

The pandemic has more negatively impacted marginalized communities, resulting in higher COVID-19 case rates and hospitalizations among people of color.^7-8^ As a result, life expectancy among Black and Hispanic adults has been reduced due to the pandemic.^9^ Earlier research addressing vaccine acceptance among racial/ethnic minorities showed lower rates of flu vaccination among Black individuals and higher levels of resistance to the flu vaccine.^10-11^ To prevent adding to healthcare disparities already highlighted during the pandemic, we need to better understand how different communities of color feel about the COVID-19 vaccine and how we can personalize vaccine campaign strategies. Black people make up just 12% of Arkansans who have received both vaccine doses, even though they account for 15% of the state’s COVID-19 deaths^12^ and 16% of the state’s population.^13^

Evidence on the distribution of COVID-19 vaccine acceptance in populations is emerging. In a sample of just over 600 American adults, more than two thirds reported they would accept a COVID-19 vaccination if recommended for them.^14^ Vaccine acceptance was measured by a single item. Results showed significant differences by gender, age, race/ethnicity, education, perception of COVID-19 risk, and region of the United States.^14^ The study, however, was limited due to sampling, which required a specific smart phone app to participate. Other studies of vaccine acceptance across the globe have generally found those who perceived themselves at higher risk and had previously been vaccinated for the influenza are more likely to be vaccinated for COVID-19.^6,14-19^ However, these studies, too, used single measures of vaccine acceptance. Only two known studies in the U.S. explicitly examined racial and ethnic differences in vaccine hesitancy in minority groups compared to non-Hispanic White adults.^20-21^ Longitudinal evidence showed the time during the pandemic when a respondent was asked about intent to get vaccinated (early/late) was associated with degree of uptake.^22-23^ Both studies were completed prior to the FDA emergency use authorization of the COVID-19 vaccines and again used single item measures.

In this paper, we examine vaccine acceptance among a random sample of Arkansas adults. Vaccine acceptance is the willingness to receive a vaccine. Unlike previous COVID-19 research, we used a newly developed, multi-component measure of vaccine acceptance.^24^ This measure was developed to address the short-comings of several vaccine hesitancy measures, including other measures principally developed to assess parents’ willingness to have their children vaccinated. The study was conducted in a small Southern rural state. This is relevant because vaccination rates are known to be lower in the rural South, where preventive medical services are generally less available.^25-27^ In this paper, we explore the utility of a multi-dimensional scale and whether using a multi-dimensional scale provides insights into health disparities in vaccine acceptance.

## Materials and Methods

### Study Population

Data were collected from 1,536 residents of Arkansas between October 3 and October 17, 2020. There was no racial or gender bias in the selection of participants. Participants were contacted using random digit dialing of mixed ground line and targeted cell phone numbers in Arkansas. Land lines were a random sample of all known land lines in Arkansas. Cell phone numbers were a random sample of active numbers used in Arkansas. Usage in the state was determined by the volume of calls and locations where a particular cell phone was used most. This target approach eliminated cell numbers not currently in use or used outside the state at the time of the poll. The mixed sample of ground and cell phones was purchased from a national company having access to these data and experience doing telephone polling in Arkansas. Samples of phone numbers were received from the company every two weeks.

Trained research assistants (RAs) called telephone numbers from a randomized non-duplicative list. If an eligible person answered the call, the RA explained that he/she was calling from a health science center and asked if the respondent was interested in answering questions about the COVID-19 pandemic. If the person refused to participate or the phone was not answered, the RA proceeded to the next number. In the event the person reached was only Spanish speaking, an RA fluent in Spanish spoke with the respondent.

After expressing willingness to participate, the RA asked if the respondent was: 18 years or older; a resident of Arkansas; able to understand and speak English or Spanish; and willing to provide informed consent. The willingness to consent and going forward with the survey was used as implicit consent. Study procedures were approved by an institutional review board for the protection of human subjects (# 260974).

### Measures

Vaccine acceptance was measured using a 10-item short version of a scale developed by Sarathchandra et al.^24^ Study participants were asked to focus on a COVID-19 vaccine when answering the questions. The scale is composed of five subscales (two items each) measuring distinct dimensions of vaccine acceptance. The five dimensions are: perceived safety of a vaccine; perceived vaccine effectiveness and necessity; acceptance of vaccine selection and scheduling of doses; perceived value of and affect toward the vaccine; and the perceived legitimacy of authorities to require people be vaccinated. Responses were recorded on a seven-point Likert-type scale ranging from one (strongly disagree) to seven (strongly agree). The mid-point of the scale was neither disagree nor agree. To arrive at a vaccine acceptance score, negatively worded questions were reverse scored, and all items were recoded to create a scale ranging from negative three (disagree) to positive three (agree) with zero as the midpoint. The overall scale’s Cronbach’s alpha for the 10 items measuring vaccine acceptance was high, 0.83.^24^ We present descriptive information on the five subscales, Cronbach’s α = 0.85 to 0.91,^24^ but only report on the overall vaccine acceptance in multivariable analyses since the short-form was used.

Study respondents were asked to provide their gender (male/female), age, race/ethnicity (schema described below), income in 2019, and Arkansas county of residence. Participants were asked several questions related to attitudes and beliefs about COVID-19, prevention behaviors, and actions taken by the state government to stop the spread of the virus in the state. Among these were: perceived chances of getting COVID-19 (no chance=0 and any chance = 1), regularly wear a face mask (no/yes), perception that face masks help to control the spread of COVID-19 (no/yes), agreement with a needed state mask order (no/yes), and attending a religious or church service in person in the past 30 days (no/yes). Respondents were also asked to rank on a scale of zero to five, with zero being least and five being strongest, their feelings about the importance of being tested, feeling safe while shopping or eating in public, and optimism of the pandemic ending soon. Finally, respondents were asked if they had health insurance (yes/no).

### Statistical Analyses

Data was collected using computer assisted telephone interviews with data stored in a Research Electronic Data Capture (RedCap) database. Stata version SE 16.1 (StataCorp LLC, College Station, TX) was used to manage and analyze data. No duplicate records were discovered in the data management process. The degree of missing data and whether data were missing at random was examined for all variables. Of the 1,536 observations, 140 observations with three or more total missing values or any one missing value on the ten-item vaccine acceptance scale were dropped. Regression diagnostics for outliers indicated 186 outliers with an influence on regression results, so a decision was made to drop observations with a residual absolute value exceeding 2.0, resulting in a final sample size of 1,164. Tests for heteroscedasticity, multicollinearity, and specification error did not reveal significant problems. To explore racial differences, we stratified the regression results for Black and White. The Hispanic and other categories were too small to allow presentation of stratified results.

To ensure the sample was representative, responses were weighted for age, sex, and White/non-White race/ethnicity. Weights were generated using a raking ratio estimation. Population percentages were generated using census data.^28^ The population weights were the ratio of percentages observed from the expected in each of the three-way cross-tabulated categories.

In preliminary analyses, distributional characteristics of variables were examined. A decision was made to collapse Pacific Islander/Marshallese (0.3%), Asian (0.6%), Native American (0.9%), and other (2%) into a single “other” category. Final racial/ethnic categories included in analyses were: White/Caucasian (75%), Black/African American (16%), Hispanic/Latino (4%), and other (5%). Age groups were 18 to 29 (15%), 30 to 39 (17%), 40 to 49 (16%), 50 to 59 (12%), 60 to 69 (19%), and 70 and above (21%).

Multivariable linear regression was used to examine the influence of sociodemographic characteristics and pandemic mitigation attitudes and behaviors on vaccine acceptance. In preliminary analyses, it was determined region of the state, optimism, and feeling safe were not statistically (p > .20) or conceptually significant and were removed from the final regression models. In final models, a conventional statistical significance level (p ≤ .05) was used.

## Results

Table 1 shows mean scores of vaccine acceptance by sociodemographic characteristics. Results indicate noticeable differences by race and income. Females were slightly more accepting of the vaccine and for perceived effectiveness, selection, and legitimacy of authorities to require vaccinations subscales. White and Hispanic respondents were more accepting compared to Black and ‘other’ respondents for each subscale. Black participants had the only mean score less than zero, which was on the vaccine safety subscale. Vaccine acceptance scores tended to increase as income increased, indicating higher income respondents were more accepting of a COVID-19 vaccine.

**Table 1:**
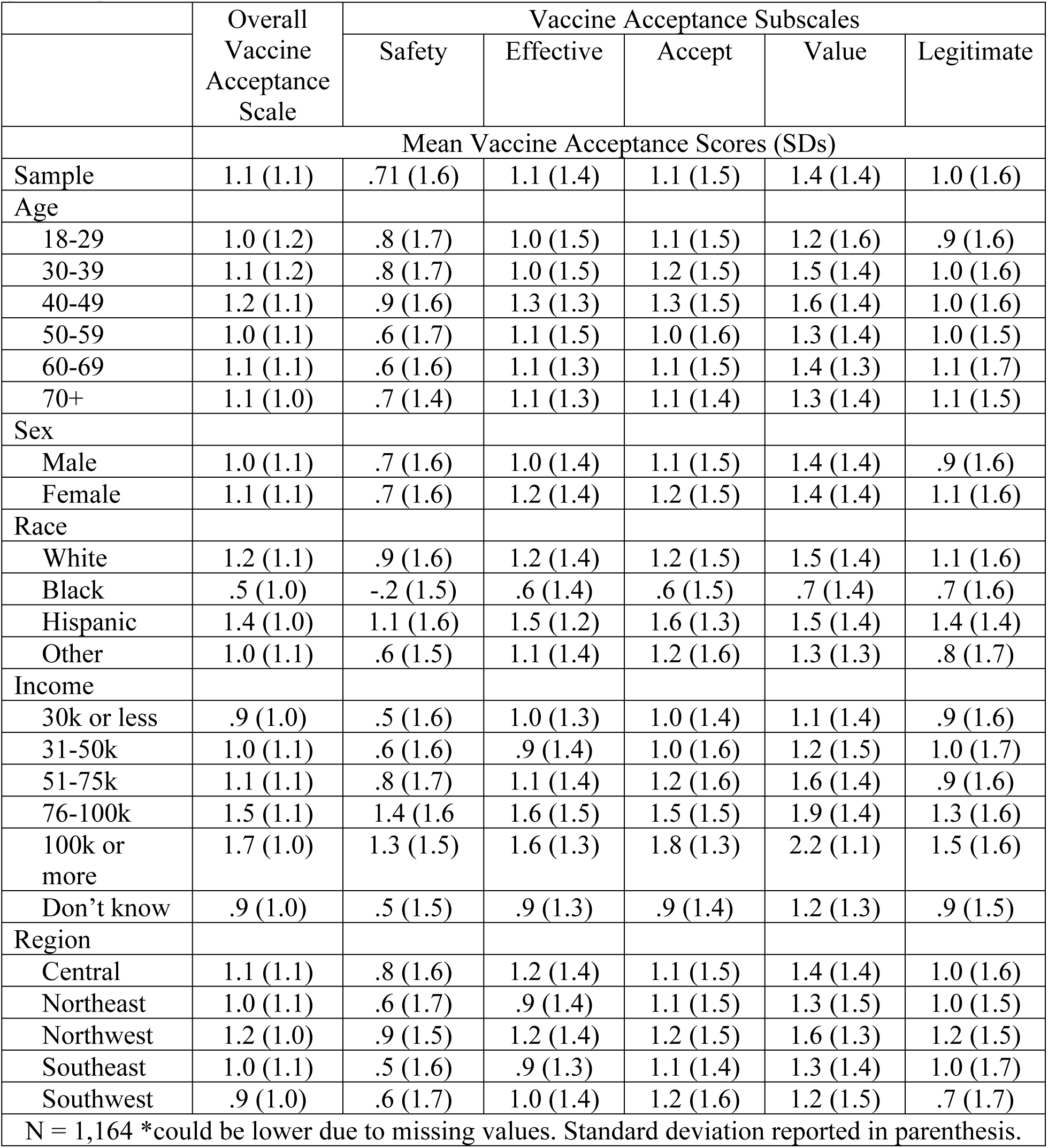
Vaccine acceptance (overall and subscales) scores by sociodemographic characteristics (n=1,164).

**Table 2:** Vaccine acceptance (overall and subscales) scores by COVID-19 attitudes, mitigation behaviors, and actions taken by the state (n=1,164).

|  | Overall<br>Vaccine<br>Acceptance<br>Scale | Vaccine Acceptance Subscales |  |  |  |  |
| --- | --- | --- | --- | --- | --- | --- |
|  |  | Safety | Effective | Accept | Value | Legitimate |
|  | Mean Vaccine Acceptance Scores (SDs) |  |  |  |  |  |
| Sample | 1.1 (1.1) | .71 (1.6) | 1.1 (1.4) | 1.1 (1.5) | 1.4 (1.4) | 1.0 (1.6) |
| Chances of<br>getting<br>COVID-19 |  |  |  |  |  |  |
| Any chance | 1.2 (1.1) | .8 (1.6) | 1.2 (1.3) | 1.2 (1.5) | 1.5 (1.4) | 1.1 (1.5) |
| No chance | .7 (1.1) | .4 (1.7) | .7 (1.5) | .9 (1.6) | 1.0 (1.5) | .6 (1.7) |
| Regular mask<br>wearer |  |  |  |  |  |  |
| Yes | 1.1 (1.1) | .8 (1.6) | 1.1 (1.4) | 1.2 (1.5) | 1.4 (1.4) | 1.1 (1.5) |
| No | .1 (1.1) | -.4 (1.8) | .4 (1.3) | .4 (1.6) | .6 (1.5) | -.4 (1.7) |
| Masks help |  |  |  |  |  |  |
| Yes | 1.2 (1.0) | .9 (1.5) | 1.2 (1.3) | 1.2 (1.4) | 1.5 (1.4) | 1.2 (1.5) |
| No | .6 (1.2) | .1 (1.7) | .6 (1.4) | .8 (1.7) | 1.1 (1.5) | .3 (1.7) |
| State face<br>mask order<br>needed |  |  |  |  |  |  |
| Yes | 1.2 (1.1) | .8 (1.5) | 1.2 (1.3) | 1.2 (1.4) | 1.4 (1.4) | 1.2 (1.5) |
| No | .9 (1.1) | .5 (1.7) | .9 (1.4) | 1.0 (1.6) | 1.3 (1.5) | .7 (1.7) |
| Importance of<br>being tested |  |  |  |  |  |  |
| Important | 1.1 (1.1) | .8 (1.5) | 1.1 (1.4) | 1.2 (1.5) | 1.4 (1.4) | 1.1 (1.5) |
| Not<br>important | .8 (1.2) | .3 (1.8) | .9 (1.4) | .9 (1.6) | 1.2 (1.4) | .5 (1.7) |
| Feel safe<br>shopping /<br>eating out |  |  |  |  |  |  |
| Safe | 1.0 (1.1) | .7 (1.6) | 1.0 (1.4) | 1.1 (1.5) | 1.4 (1.4) | .9 (1.6) |
| Unsafe | 1.2 (1.1) | .7 (1.6) | 1.2 (1.3) | 1.3 (1.4) | 1.4 (1.4) | 1.2 (1.5) |
| Optimism<br>pandemic will<br>end soon |  |  |  |  |  |  |
| Optimistic | 1.0 (1.1) | .6 (1.7) | 1.0 (1.4) | 1.1 (1.5) | 1.2 (1.4) | .8 (1.7) |
| Not<br>optimistic | 1.2 (1.1) | .8 (1.6) | 1.2 (1.4) | 1.2 (1.5) | 1.5 (1.4) | 1.2 (1.5) |
| Attend social<br>gatherings |  |  |  |  |  |  |
| Yes | 1.1 (1.2) | .7 (1.7) | 1.1 (1.4) | 1.2 (1.5) | 1.5 (1.4) | .9 (1.7) |
| No | 1.1 (1.1) | .7 (1.6) | 1.1 (1.4) | 1.1 (1.5) | 1.4 (1.4) | 1.1 (1.6) |
| Attend religious services |  |  |  |  |  |  |
| Yes | .9 (1.2) | .5 (1.7) | 1.0 (1.4) | 1.1 (1.6) | 1.3 (1.5) | .8 (1.7) |
| No | 1.1 (1.1) | .8 (1.6) | 1.1 (1.4) | 1.2 (1.5) | 1.4 (1.4) | 1.1 (1.6) |
| Health insurance |  |  |  |  |  |  |
| Yes | 1.1 (1.1) | .8 (1.6) | 1.1 (1.4) | 1.2 (1.5) | 1.4 (1.4) | 1.1 (1.6) |
| No | .6 (1.3) | .1 (1.7) | .6 (1.5) | .8 (1.8) | 1.0 (1.6) | .4 (1.8) |
| N = 1,164 *could be lower due to missing values. Standard deviation reported in parenthesis. |  |  |  |  |  |  |

Results from multivariable linear regression models indicated Black participants (p ≤ .001) were significantly less accepting of a COVID-19 vaccine, while Hispanic participants (p = .02) were more accepting, as shown in Table 3. Those with an income of more than $51,000 were more accepting than those with incomes less than $30,000. Several COVID-19 attitudes and behaviors had a significant positive association with vaccine acceptance: perceived chances of getting COVID-19 (p = .04), regularly wearing a mask (p ≤ .001), belief that masks help stop the spread of COVID-19 (p ≤ .001), believed being tested is important (p = .02), and had health insurance (p = .04). Attending church or religious services in person (p = .02) was the only factor showing a negative association with vaccine acceptance.

**Table 3:** Regression of vaccine acceptance on sociodemographic characteristics and COVID-19 attitudes and behaviors for the overall population and for Black and White adults.

| Variable | Overall<br>(n=1,164) | White<br>(n=876) | Black<br>(n=187) |
| --- | --- | --- | --- |
| Intercept | -1.03 (.22) | -1.23 (.25) | -.34 (1.16) |
| Age | - | - | - |
| 18-29 | - | - | - |
| 30-39 | .01 (.10) | -.06 (.12) | .22 (.24) |
| 40-49 | .11 (.10) | .08 (.12) | .12 (.24) |
| 50-59 | -.03 (.11) | -.05 (.14) | .25 (.25) |
| 60-69 | -.04 (.10) | -.10 (.12) | .27 (.24) |
| 70+ | .00 (.10) | -.04 (.11) | .35 (.25) |
| Sex | - | - | - |
| Male | - | - | - |
| Female | .09 (.06) | .03 (.07) | .48** (.14) |
| Race | - | - | - |
| White | - | - | - |
| Black | -.81** (.08) | - | - |
| Hispanic | .35* (.15) | - | - |
| Other | -.16 (.13) | - | - |
| Income | - | - | - |
| 30k or less | - | - | - |
| 31-50k | .18 (.10) | .16 (.11) | .33 (.20) |
| 51-75k | .24* (.10) | .23* (.12) | .22 (.25) |
| 76-100k | .71** (.11) | .76** (.13) | .24 (.32) |
| 100k or more | .85** (.11) | .86** (.13) | .87** (.27) |
| Don't know | .01 (.08) | .01 (.09) | .15 (.18) |
| Any chance of getting COVID-19 | .15* (.07) | .10 (.08) | .50** (.16) |
| Regular mask wearer | .79** (.13) | .82** (.13) | -.73 (.97) |
| Masks help stop the spread | .41** (.08) | .37** (.09) | .47* (.22) |
| State face mask order needed | .10 (.07) | .10 (.08) | -.13 (.25) |
| Importance of being tested | .18* (.08) | .25** (.09) | -.23 (.30) |
| Attends social gatherings | .10 (.07) | .18* (.08) | -.22 (.19) |
| Attends religious services | -.16* (.07) | -.21** (.08) | .09 (.17) |
| Has health insurance | .42** (.12) | .61** (.15) | .63* (.30) |
|  | Adj R<sup>2</sup> = .25 | Adj R<sup>2</sup> = .22 | Adj R<sup>2</sup> = .18 |
| Standard errors are reported in parentheses.<br>*/** Indicates significance at the .05/.01 level. |  |  |  |

The effects by race/ethnicity are shown in Table 3. Being a Black woman had a significant positive association with vaccine acceptance (p ≤ .001). Having an income above $50,000 had a positive relationship for White participants, and more than $100,000 for Black participants. Significant racial differences were noted in COVID-19 attitudes and behaviors. Regularly wearing a face mask (p ≤ .001), belief that masks help stop the spread of COVID-19 (p ≤ .001), belief that it was important to get tested for COVID-19 (p ≤ .001), and having health insurance (p = ≤ .001) in White participants was found to be significantly related to greater vaccine acceptance. Attending church or a religious service (p = .01) had a negative relationship on vaccine acceptance for White participants. Belief that masks help stop the spread of COVID-19 (p = .03), having health insurance (p = .04), and higher perceived chances of getting COVID-19 (p ≤ .01) were related to vaccine acceptance among Black participants.

## Discussion

In a representative sample of Arkansans, we found relatively tepid acceptance of a COVID-19 vaccine in the period just proceeding the emergency approval of the vaccine. Vaccine acceptance varied by race/ethnicity, income, having insurance, and beliefs and attitudes about COVID-19 mitigation behaviors. These suggest, except as noted below, it is quite likely education and income play a significant role in determining COVID-19 vaccine acceptance. This may be due, as some have suggested, to a basic understanding of and trust in science, rather than acceptance of a COVID-19 vaccine as such.^29-30^

Black Arkansans had the lowest levels of vaccine acceptance. This is true for the overall vaccine acceptance scale and for each subscale. Lower COVID-19 vaccine acceptance among Black participants may be due to mistrust in the medical establishment and continuing lack of equality in medical services throughout the South.^3^ Hispanic and White participants had higher and similar levels of acceptance. These findings strongly suggest a single vaccine acceptance campaign strategy is unlikely to convince large numbers of Black adults to be vaccinated. DeRoo et al.^3^ advocate for mobilizing minority community leaders to help motivate minority communities to be vaccinated. In the South, this certainly includes recruiting faith leaders to help motivate their congregations. Distributing the vaccine in churches after services may greatly reduce the costs of visiting a clinic for those dependent on hourly jobs. A key finding in our work is the higher positive acceptance of vaccines among Black women. If one goal is to influence social norms in a community, then relying on Black women, who appear to have more positive attitudes toward the vaccine than Black men, as community opinion leaders may be important in the Black community.

One reason this study was conducted was to examine the value of a more robust conceptualization of vaccine acceptance. The idea of a multi-dimensional scale is useful if it contributes meaningful variability. The sample is too small to conduct the robust comparisons across subscales, but the measure provided meaningful variability. Only Black participants had a negative valence on the safety subscale. No other subscale produced a negative valence. Hispanic participants have higher levels of vaccine acceptance, higher than White participants on some subscales. We believe this approach to measuring vaccine acceptance or hesitancy is an improvement over a single item measure. More psychometric research on the measure should be conducted.

This study has some limitations which should be noted. First, the sample includes the population of only one state. Therefore, findings are generalizable only within Arkansas. Even so, given similarities with the populations of other Southern states, our findings are likely more broadly indicative of COVID-19 vaccine acceptance in the South. Although non-response rates were reduced and survey results were weighted, response bias can still impact interpretations due to response patterns. Additionally, the survey was conducted just before the FDA approved the two mRNA vaccines currently available in the U.S. While the media were reporting the imminent emergency approval of the vaccines while the survey was ongoing, we were unable to assess the acceptance after formal approval of the vaccines.

## Conclusion

There is considerable work to be done to convince a sizable proportion of the population in many Southern states to accept a COVID-19 vaccine. This research shows, depending on the racial/ethnic group, there are different attitudes about a COVID-19 vaccine. Vaccine acceptance is multi-dimensional and we should avoid single item measures of acceptance. By using single item measures, we may make a conclusion, but may be unable to provide an explanation based on valid measurement. We also run the risk of aggregating different populations into a single category, thereby mischaracterizing important differences among groups that can otherwise help personalize public health messaging.

## Data Availability

All analytic de-identified files are available from REDCap (Research Electronic Data Capture)

## Author Contributions

**Benjamin C. Amick**: Conceptualization, Methodology, Investigation, Writing - Original Draft, Writing - Review & Editing, Project administration, Funding acquisition; **Jaimi L. Allen**: Conceptualization, Methodology, Formal analysis, Data curation, Writing - Original Draft, Writing - Review & Editing; **Clare C. Brown**: Conceptualization, Methodology, Formal analysis, Writing - Review & Editing; **Anthony Goudie**: Conceptualization, Methodology, Writing - Review & Editing; **Mick Tilford**: Conceptualization, Methodology, Writing - Review & Editing; **Mark Williams**: Conceptualization, Methodology, Writing - Review & Editing, Funding acquisition.

## Data availability statement

The data that support the findings will be provided upon request.

## References

1. Centers for Disease Control and Prevention. COVID Data Tracker. https://covid.cdc.gov/covid-data-tracker. Accessed July 27, 2021.

2. U.S. Department of Health & Human Services. COVID-19 State Profile Report – Arkansas. https://healthdata.gov/Community/COVID-19-State-Profile-Report-Arkansas/cdsu-kww8. Accessed July 27, 2021.

3. DeRoo S, Pudalov N, & Fu L. Planning for a COVID-19 vaccination program. JAMA. 2020;323(24):2458–2459. doi:10.1001/jama.2020.8711

4. Gostin L & Salmon D. The dual epidemics of COVID-19 and influenza: Vaccine acceptance, coverage, and mandates. JAMA. 2020;324(4):335–336. doi:10.1001/jama.2020.10802

5. Ozawa S. & Stack M. (2013). Public trust and vaccine acceptance-international perspectives. Hum Vaccin Immunother. 2013;9(8):1774–8. doi:10.4161/hv.24961

6. Palamenghi L, Barello S, Boccia S, Graffigna G. Mistrust in biomedical research and vaccine hesitancy: the forefront challenge in the battle against COVID-19 in Italy. Eur J Epidemiol. 2020;35(8):785–788. doi:10.1007/s10654-020-00675-8

7. Price-Haywood EG, Burton J, Fort D, Seoane L. Hospitalization and Mortality among Black Patients and White Patients with Covid-19. N Engl J Med. 2020;382(26):2534–2543. doi:10.1056/NEJMsa2011686

8. Millett GA, Jones AT, Benkeser D, et al. Assessing differential impacts of COVID-19 on black communities. Ann Epidemiol. 2020;47:37–44. doi:10.1016/j.annepidem.2020.05.003

9. Andrasfay T and Goldman N. Reductions in 2020 US life Expectancy due to COVID-19 and the disproportionate impact on Black and Latino populations. PNAS. 2021; 118(5) 1–6. doi: 10.1073/pnas.2014746118

10. Hebert PL, Frick KD, Kane RL, McBean AM. The causes of racial and ethnic differences in influenza vaccination rates among elderly Medicare beneficiaries. Health Serv Res. 2005;40(2):517–537. doi:10.1111/j.1475-6773.2005.00370.x

11. Quinn SC, Lama Y, Jamison A, Freimuth V, Shah V. Willingness of Black and White Adults to Accept Vaccines in Development: An Exploratory Study Using National Survey Data. American Journal of Health Promotion. 2021;35(4):571–579. doi:10.1177/0890117120979918

12. Arkansas Department of Health. COVID-19 Dashboard. https://experience.arcgis.com/experience/633006d0782b4544bd5113a314f6268a/page/page_0/. Accessed July 27, 2021.

13. United States Census Bureau. QuickFacts: Arkansas. https://www.census.gov/quickfacts/AR. Accessed July 27, 2021.

14. Malik A, McFadden S, Elharake J, & Omer S. Determinants of COVID-19 vaccine acceptance in the U.S. EClinicalMedicine. 2020;26. doi:10.1016/j.eclinm.2020.100495

15. Dror A, Eisenbach N, Taiber S, Morozov N, Mizrahi M, Zigron A, Srouji S, & Sela E. Vaccine hesitancy: The next challenge in the fight against Covid-19. Eur J Epidemiol. 2020;35,775–779. doi:10.1007/s10654-020-00671-y

16. Fisher K, Bloomstone S, Walder J, Crawford S, Fouayzi H, Mazor K. Attitudes toward a potential SARS-CoV-2 Vaccine: A survey of U.S. adults. Ann Intern Med. 2020;173(12):964–973. doi:10.7326/M20-3569.

17. Lazarus J, Ratzan S, Palayew A, Gostin L, Larson H, Rabin K, Kimball S, & El-Mohandes A. A global survey of potential acceptance of a COVID-19 vaccine. Nat Med. 2021;27,225–228. doi:10.1038/s41591-020-1124-9

18. Pogue K, Jensen JL, Stancil CK, et al. Influences on Attitudes Regarding Potential COVID-19 Vaccination in the United States. Vaccines. 2020;8(4):582. doi:10.3390/vaccines8040582

19. Wang J, Jing R, Lai X, et al. Acceptance of COVID-19 Vaccination during the COVID-19 Pandemic in China. Vaccines. 2020;8(3):482. doi:10.3390/vaccines8030482

20. Willis DE, Andersen JA, Bryant-Moore K, et al. COVID-19 vaccine hesitancy: Race/ethnicity, trust, and fear. Clin Transl Sci. 2021;00:1–8. doi: 10.1111/cts.13077

21. Nguyen LH, Joshi AD, Drew DA, et al. Racial and ethnic differences in COVID-19 vaccine hesitancy and uptake. medRxiv. 2021;2021.02.25.21252402. doi:10.1101/2021.02.25.21252402

22. Daly M and Robinson E. Willingness to vaccinate against COVID-19 in the US: Longitudinal evidence from a nationally representative sample of adults from April-October 2020. MedRxiv (2020). http://doi.org/10.110/202011.27.20239970.

23. Fridman A, Gershon R, Gneezy A. COVID-19 and vaccine hesitancy: A longitudinal study. Plos One (2021) 16(4): e0250123. Doi.org/10.1371/.

24. Sarathchandra D, Navin MC, Largent MA, McCright AM. A survey instrument for measuring vaccine acceptance. Prev Med. 2018;109:1–7. doi:10.1016/j.ypmed.2018.01.006

25. Casey MM, Thiede Call K, Klingner JM. Are rural residents less likely to obtain recommended preventive healthcare services?. Am J Prev Med. 2001;21(3):182–188. doi:10.1016/s0749-3797(01)00349-x

26. Chui A, Dushoff J, Yu D, & He D. Patterns of influenza vaccination coverage in the United States from 2009 to 2015. Int J Infect Dis. 2017;65,122–127. doi:10.1016/j.ijid.2017.10.004

27. Zhai Y, et al. Rural, urban, and suburban differences in influenza vaccination coverage among children. Vaccine. 2020;38, 7596–7602. doi:10.1016/j.vaccine.2020.10.030

28. Arkansas Economic Development Institute. Population estimates & projections. https://arstatedatacenter.youraedi.com/population-estimates-projections/. Accessed January, 2021.

29. Romer D, Jamieson KH. Conspiracy theories as barriers to controlling the spread of COVID-19 in the U.S. Soc Sci Med. 2020;263:113356. doi:10.1016/j.socscimed.2020.113356

30. SteelFisher GK, Blendon RJ, & Caporello H. An Uncertain Public-Encouraging Acceptance of Covid-19 Vaccines. N Engl J Med. 2021;384:1483–1487. doi:10.1056/NEJMp2100351

